# Effort-based Decision-Making in Post-Stroke Gait: A Feasibility Study

**DOI:** 10.64898/2026.01.28.26344556

**Authors:** James Sulzer, Dana Lorenz, Ben Killen, Jake Stahl, Abigail Farrell, Sophie Osada, Mark Waschak, Vikram Chib, Michael Lewek

## Abstract

Conventional therapy after stroke focuses on reducing physical impairments. However, the decisions that guide people’s movements may have far-reaching consequences towards recovery. We lack the tools to characterize these decisions. Recently, researchers have created a quantitative behavioral assessment of effort-based decision-making and applied it to some clinical populations. The purpose of this paper is to examine the feasibility of evaluating effort-based decision-making during walking after stroke. We recruited five neurotypical participants in an initial study. We conducted a subjective effort valuation on the neurotypical individuals with and without a knee immobilizer to simulate the biomechanics of reduced knee flexion during post-stroke gait. Participants cleared obstacles of varying heights during overground walking, followed by rating their perceived effort and then completing an effort choice paradigm to calculate subjective effort value. In a second experiment, we recruited five individuals with stroke to perform a similar protocol without an immobilizer during harnessed treadmill walking. We found that rated perceived effort increased monotonically with obstacle height across groups, that individuals could recall obstacle heights without cues, and that subjective effort value increased with knee immobilization in the control group as expected. We conclude that adapting an effort-based decision-making assessment to a walking context in people with stroke is feasible.

## I. Introduction

Mobility and walking are highly influential in maintaining quality of life after neurological injury such as stroke [1], [2], [3]. People with post-stroke Stiff-Knee gait (SKG), defined as reduced paretic peak knee flexion during swing phase, have difficulty during walking, particularly in maintaining foot clearance [4]. The reduced foot clearance can result in energy-consuming gait compensations and lead to higher risk of falls [5], [6], [7]. Although these consequences of altered gait can discourage mobility, actively challenging oneself and maintaining effort is critical to driving recovery [8]. Thus, while the individual may understand *how* to achieve a task, it is equally as important to have the willingness to exert the effort required to complete the activity. These effort-based movement decisions influence daily activity and function. Despite the clear importance of psychological contributors to recovery [9], rehabilitation practice almost entirely focuses on physical impairments. Our goal is to quantitatively investigate how psychological factors, namely effort-based decision-making, influence activity and recovery after stroke.

Effort-based decision-making has been quantified using principles from behavioral economics. Movement decisions are a consequence of weighing the required effort and the anticipated reward resulting from action [10], [11], [12]. The perceived effort involved with a motor task is derived from both physical exertion and mental effort [13], [14]. To isolate the effect of expected effort, a novel approach to investigate physical exertion in decision-making was developed as a forced choice between two alternatives [15]. In this approach, participants choose between a certain outcome representing a given level of effort (for example, squeezing a hand grip at 30% maximum voluntary force) versus an uncertain (gamble) outcome (for example, a 50% chance of squeezing at 60% maximum voluntary force, or a 50% chance of no required squeeze at all). Assuming a linear relationship between perceived effort and grip force, these alternatives should have equivalent expected values. In other words, the prospect of a 50% chance of exerting twice the effort of the alternative at 100% probability should produce the same expected value.

The willingness to exert effort is variable between individuals and is known as the subjective effort value [14], [15], [16], [17]. Some individuals are naturally more willing to exert effort (placing a low subjective effort value on a task) and others are less willing (placing a higher subjective effort value on an action). These differences in subjective effort valuation have been linked to neural substrates such as the ventromedial prefrontal cortex [15]. Subjective effort value emerges as a result of multiple physiological and psychological factors, including motivation, fatigue and pain [14], [16]. Alterations in effort-based decision-making behavior have been found in psychological and psychiatric conditions such as chronic fatigue, anxiety, depression, schizophrenia, and attention disorders [18]. Critically, many of these disorders co-exist after stroke [19].

The overall objective of this work is to determine the feasibility of translating the behavioral economics approach of obtaining subjective effort value to walking in individuals post-stroke. We conducted two experiments to determine feasibility. The first goal was to evaluate the feasibility of translating the evaluation of subjective effort to a walking task. Our hypothesis was that subjective effort value would increase with increasing task difficulty. We increased task complexity by altering gait biomechanics using a knee immobilizer in neurotypical participants to mimic gait patterns consistent with post-stroke SKG [5], [20]. The second experiment translated this procedure to individuals with post-stroke SKG. Given the additional confound of neuromotor coordination deficits post-stroke, we hypothesized that subjective effort values obtained in individuals post-stroke would be greater than those in neurotypical individuals with knee immobilization. Feasibility was assessed by the consistency and monotonicity of the ratings of perceived effort. This work is a novel adaptation of effort-based decision-making to sensorimotor rehabilitation.

## II. Experment 1

Investigating subjective effort value requires a motor task with a parametric modulation of difficulty. This has typically been investigated using button presses or precision grip tasks of different isometric force levels [15], [21]. However, these tasks do not reflect the gait-based decision making that is important for people with SKG. Instead, our proposed work uses obstacle clearance, which: 1) targets the main deficit of people with SKG (i.e., foot clearance); 2) can be easily modulated (change in obstacle height); 3) is expected to increase effort ratings monotonically as obstacle height increase, unlike variance in gait speed[22]; 4) is not as fatiguing as walking faster or for longer bouts; and 5) is a clinically relevant predictor of falls [23].

We recruited 5 neurotypical individuals, ages 24-29 (2 M / 3 F), with heights ranging from 1.65-1.78m, approved by the institutional review board at the University of North Carolina – Chapel Hill. The experimental setup consisted of a 20-foot walkway with four adjustable-height obstacles spaced four feet apart. Markerless motion capture (OpenCap) was used to monitor kinematics. Knee immobilization was achieved using a three-panel knee immobilizer with reinforced medial, lateral, and posterior stays to prevent knee flexion (Hely & Weber Model 214). Our earlier work showed that the use of knee immobilization produced knee flexion angles equivalent to people with SKG post-stroke[5], [24]. For each individual, we measured the maximum foot clearance height while standing.

The experiment occurred over a single day. The first phase was called the *Association* phase. The purpose of the Association phase was for participants to build an association between the external stimulus (obstacle height) and rating of perceived effort (RPE). Participants were asked to walk at a comfortable speed across a level walkway to obtain baseline gait parameters (Fig. 1). The participant was then exposed to 24 trials of walking over obstacles, with each of the six heights presented four times and ordered pseudorandomly in blocks of two. This was then repeated to allow participants to experience all of the obstacles so that they could contextualize the effort required for each obstacle. Each of the six obstacle heights were presented as its own color (linearly ranging from purple = 1” to red = 100% maximum clearance) (Fig. 1). After each pair of trials, we asked the participant to rate their perceived effort on a scale of 1 to 10, with 10 being maximum effort (or the highest obstacle). Instances where the hurdles were bumped or knocked over were recorded. Participants were asked to match their comfortable overground gait speed as closely as possible and received feedback on the time each pass took. This phase lasted about 20 minutes.

**Fig 1.**
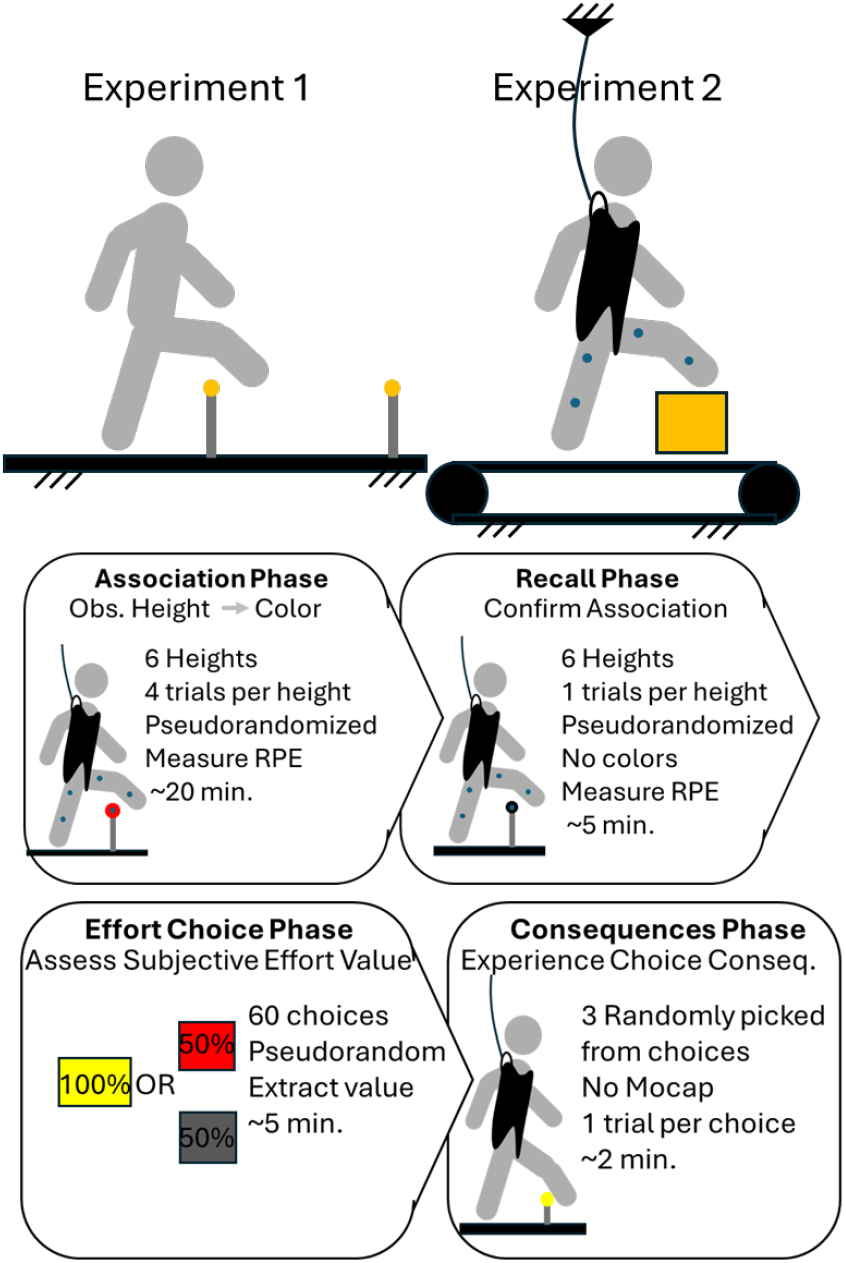
In Experiment 1, neurotypical participants walk overground over obstacles and repeat the association, effort choice and consequences phases with knee immobilization. In Experiment 2, participants with stroke walk over foam blocks on a treadmill with fall arrest support. Below, a summary of the protocol, with association, recall, effort choice and consequence phases.

The second phase was the *Recall* phase. The purpose of the recall phase was to assess the extent to which the participant has internalized the association between the height of the obstacle and RPE. In this phase, participants complete one trial for each of the previously associated heights in pseudorandomized order while attempting to match their comfortable gait speed, but all obstacles were colored white. Participants then stated their RPE and guessed the associated color for that obstacle height. This phase lasted about 5 minutes.

The third phase was the *Effort choice* phase. The purpose of the Effort choice phase was to measure how participants value effort exertion. Each participant answered 60 effort choice questions on a desktop computer, choosing between a ‘sure’ effort level, or a ‘gamble’ effort level. The participant may select to gamble, in which their choice amounts to a “coin flip” certainty, consisting of either a high obstacle height or no obstacle at all. Alternatively, the participant may select a ‘sure’ option, consisting of an obstacle at a lower height. The height values vary across conditions, but the “gamble” (uncertain) condition is always the higher height. This phase lasted about 5 minutes.

The fourth phase was the *Consequences* phase. The purpose of the Consequences phase was to complete a subset of the choices made in the effort choice phase. In this last phase, the participant completed one walking trial for three of the obstacle choices made at random during the effort choice phase. Participants were instructed earlier that they would not be able to leave the testing area until they successfully completed each obstacle. This phase lasted about 2 minutes.

We then completed the Association phase again, but this time with participants wearing the knee immobilizer and performing a single repetition of the phase. After this second Association phase, we ran a second Effort choice phase and then a final Consequences phase, altogether lasting about another 25 minutes.

Our goal was to determine feasibility, which means that 1) RPE monotonically increases with obstacle height, 2) participants can repeatably perceive effort without color cues for obstacle heights, 3) subjective effort values could be increased with increased task difficulty. We extracted the effort choice data and fit a logistic regression curve across the range of expected value differences between conditions using custom Matlab code. Our main performance outcome measures were RPE and the subjective effort value. We calculated subjective effort value in two ways. The first was the probability of choosing the sure condition at zero relative value, which we call the *sure bias*. The second method of measuring subjective effort value was the area under the logistic regression curve above the horizontal axis, or in other words, the integrated probability of choosing the sure option, normalized to the domain of expected values. This is the *valuation integral*.

To compare between conditions, we used a Wilcoxon signed-rank test (a<0.05). We expected knee immobilization would increase these parameters due to the higher effort needed to clear obstacles.

## III. Experment 2

We ran a separate but related experiment, recruiting 5 individuals with post-stroke SKG. Inclusion criteria were the ability to provide informed consent and follow instructions, over 18 years of age, able to lift the foot in standing to clear a 4” obstacle, premorbidly independent ambulator, presence of SKG as determined by a clinician (i.e. reduced swing phase knee flexion relative to non-paretic limb, absent of genu recurvatum), and present hemiparesis. We excluded any individuals with stroke if they had multiple strokes, sub-tentorial lesions, concomitant neurological diagnosis, lower limb joint pain, functionally relevant cognitive impairment, or fixed equinus deformity. Participants provided informed consent according to the Institutional Review Board at MetroHealth Medical Center. Participant data is shown in Table 1. As in Experiment 1, we measured the maximum clearance height of the paretic foot.

**Table 1:** Characteristics of participants with stroke.

| ID | Age/Sex | Height | Side of stroke | Time since stroke (yrs) | Comf. gait speed (m/s) | Clear. Height (m) |
| --- | --- | --- | --- | --- | --- | --- |
| S1 | 60's/M | 1.88 | R | 10 | 0.26 | 0.15 |
| S2 | 70's/M | 1.80 | L | 11 | 0.32 | 0.22 |
| S3 | 70's/F | 1.80 | R | 12 | 0.24 | 0.25 |
| S4 | 50's/M | 1.83 | R | 4 | 0.38 | 0.46 |
| S5 | 60's/M | 1.93 | R | 8 | 0.15 | 0.17 |

Instead of the overground walking scenario in Experiment 1, in Experiment 2 participants walked on a split-belt treadmill (Bertec, Columbus, OH) while kinematics were recorded using optical motion capture (Vicon, Oxford, UK), with a standard lower body marker set. For obstacles, we used 12” deep colored foam blocks that extended across the width of the treadmill (Fig. 1). The height range was linearly distributed from 20%-100% of the maximum clearance using the same color scheme as in Experiment 1. The blocks were cut to heights based on different intervals, with some blocks acting as shims to match the exact height specifications. To protect from falls, we used the Bertec overhead harness. Handrails were available for participants to use in lieu of an assistive device such as a cane. All participants who used ankle-foot orthoses continued to use them in the experiment.

We used the same subjective effort valuation protocol as in Experiment 1. Instead of overground walking, participants walked on the treadmill at 70% of their comfortable gait speed. The foam obstacles were placed in front of the participant by one of the researchers and then collected by another researcher after the participant cleared the obstacle. We also employed the same data management steps as in Experiment 1. We compared people with SKG to neurotypical controls walking with and without the knee immobilizer using a Wilcoxon rank sum test (a<0.05).

## IV. Results

All 10 participants across both sites completed the experiment with no adverse events.

One measure of feasibility is a monotonic increase in RPE as obstacle height increases. Fig. 2 illustrates the relationship between these parameters in both neurotypical (top) and post-stroke (bottom) individuals. There were no instances of non-monotonic changes in neurotypical individuals. Out of the six conditions and five participants with stroke (30 datapoints) we found four instances of non-monotonicity (13%), one instance distributed across four different participants.

**Fig 2.**
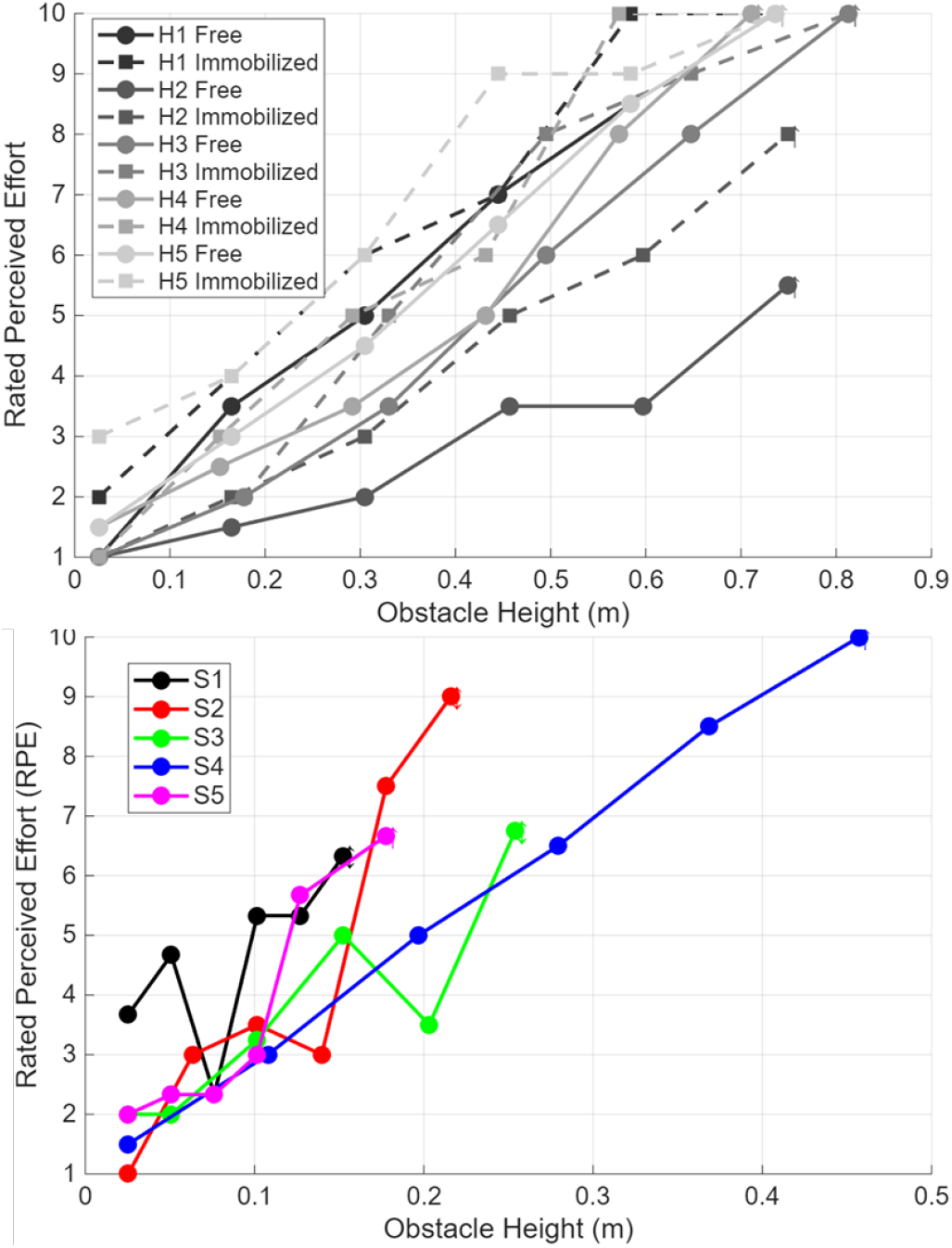
Monotonicity of rated perceived effort (RPE) during association phase compared to obstacle height in people neurotypical (H, above) and people with stroke (S, below). With or without immobilization, RPE increases with height, with some exceptions.

Another measure of feasibility is the repeatability between RPE during association and recall phases. Fig. 3 shows the correlation between the RPE during the association phase and during the recall phase. Neurotypical participants largely showed excellent recall (Fig. 3, top), whereas post-stroke participants had lower R^2^ values (Fig. 3, bottom). While most relationships were highly correlated, one post-stroke participant showed a moderate correlation (R^2^=0.55) and one neurotypical participant had only a good correlation (R^2^=0.77).

**Fig 3.**
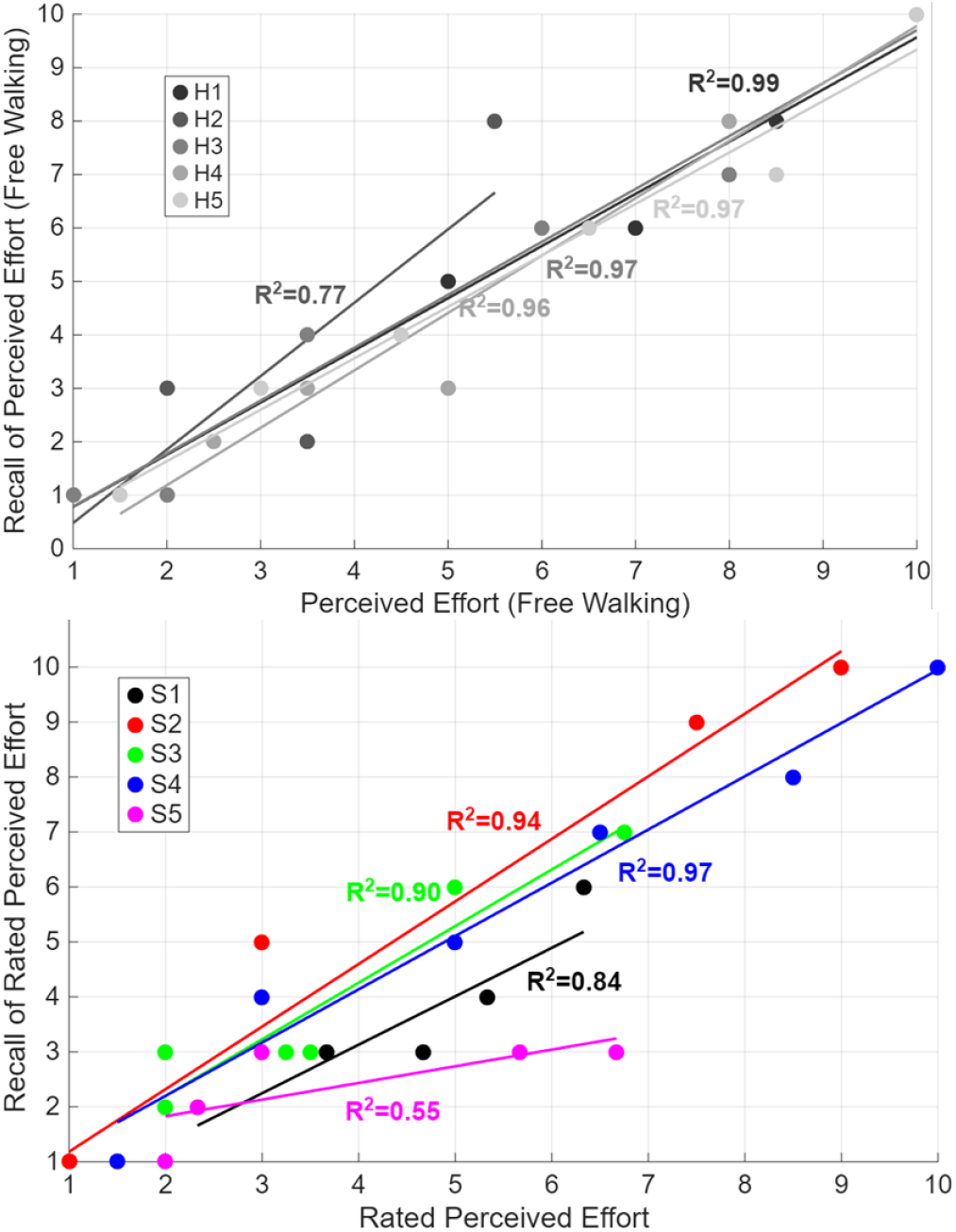
Repeatability of RPE between association and recall phases in neurotypical (H, above) and people post-stroke (S, below). Both plots show largely linear relationships between RPE and recall, but each group has one individual with lower correlation.

In addition to repeatability of RPE, we also wanted to know how well individuals could recall the specific color cue associated with the obstacle height. Table 2 shows recall scores for each participant for both groups in each condition (CR).

**Table 2:**
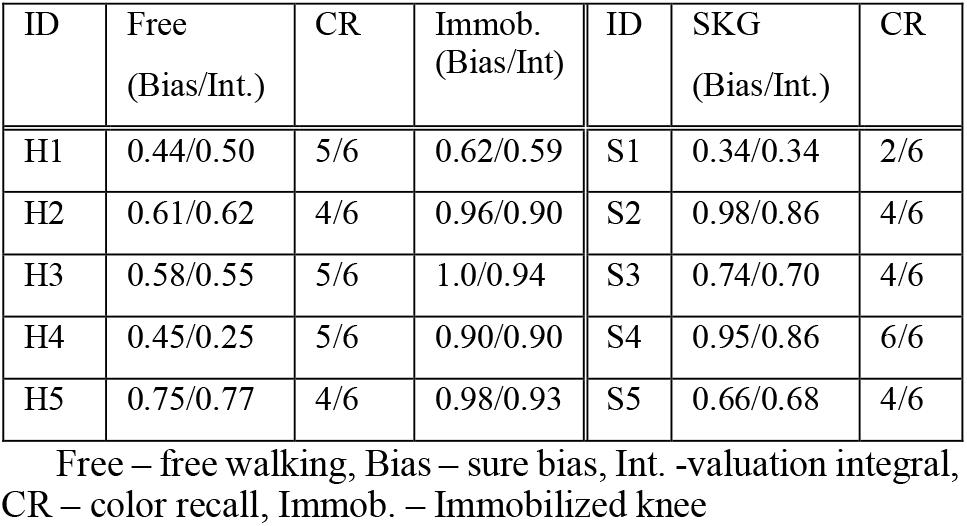
Performance metrics.

| ID | Free<br>(Bias/Int.) | CR | Immob.<br>(Bias/Int) | ID | SKG<br>(Bias/Int.) | CR |
| --- | --- | --- | --- | --- | --- | --- |
| H1 | 0.44/0.50 | 5/6 | 0.62/0.59 | S1 | 0.34/0.34 | 2/6 |
| H2 | 0.61/0.62 | 4/6 | 0.96/0.90 | S2 | 0.98/0.86 | 4/6 |
| H3 | 0.58/0.55 | 5/6 | 1.0/0.94 | S3 | 0.74/0.70 | 4/6 |
| H4 | 0.45/0.25 | 5/6 | 0.90/0.90 | S4 | 0.95/0.86 | 6/6 |
| H5 | 0.75/0.77 | 4/6 | 0.98/0.93 | S5 | 0.66/0.68 | 4/6 |

An example of a logistic regression curve and the subsequent subjective effort valuation outcomes are shown in Fig. 4. The curve shows an example during free walking where the weighting between sure and gamble conditions are approximately equal.

**Fig 4.**
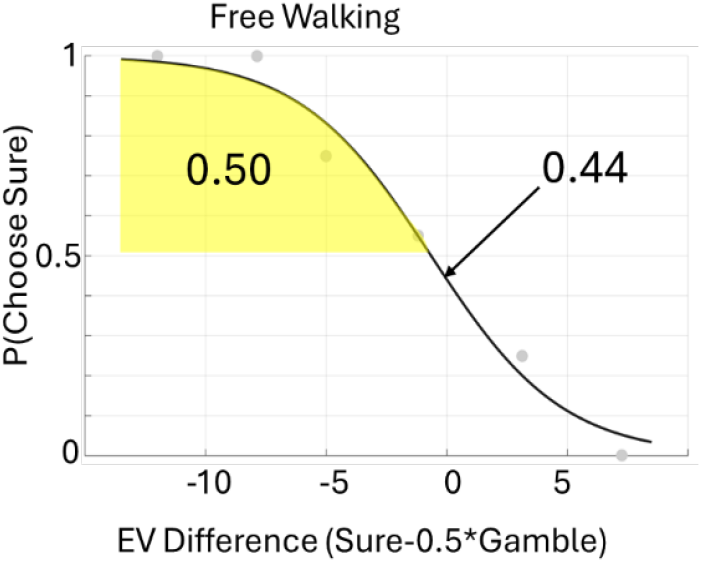
Example logistic regression curve extracted from effort choice data across domain of expected value (EV) differences. Yellow shaded area is the valuation integral (0.50), and the bias parameter is 0.44, both indicating approximately equal weighting of sure and gamble conditions in this example of free walking.

Table 2 illustrates outcome measures from both experiments. Both bias and integral scores are shown for all conditions. CR represents the color recall accuracy from the recall phase.

Free – free walking, Bias – sure bias, Int. -valuation integral, CR – color recall, Immob. – Immobilized knee Comparing free walking vs. immobilized knee walking using a one-tailed Wilcoxon signed rank test results in significant increase in sure bias (p=0.03) and valuation integral (p= 0.03).

Table 2 illustrates subjective effort valuation parameters for Experiment 2. Using a two-sided Wilcoxon rank sum test, we did not find any differences in either outcome between people with SKG and free walking or knee immobilization (p>0.05).

## V. Discussion

The goal of this study was to evaluate the feasibility of translating subjective effort valuation to a walking task in neurotypical individuals and people with post-stroke SKG. We evaluated the degree to which RPE increased monotonically with obstacle height, repeatability of RPE without color cues, color recall accuracy and two different measures of subjective effort value. We found that participants could complete the protocol successfully without adverse events, evidence of monotonic relationships for RPE with obstacle height, good recall, and expected ability to modulate subjective effort value with knee immobilization. This is the first adaptation of this paradigm to walking and the first time this type of paradigm was used in people after stroke. Understanding how subjective valuation of effort affects motor behavior could help rehabilitation researchers and providers leverage behavioral economics approaches to better personalize interventions for individuals with post-stroke motor impairment.

RPE increased monotonically with obstacle height, though some exceptions occurred. In individuals post-stroke, we found four instances where RPE decreased out of 30 instances (Fig. 2). There was no apparent relation of these decreases to height or color, and all were lower RPEs than expected. It is possible that these simply represented natural variability in performance over time [25]. Increased variability in RPE and performance could also have influenced the repeatability of RPE shown in Fig. 3.

The ability to recall the condition without the color cue likely influences the individual’s ability to make accurate effort choices. We found approximately similar recall results between neurotypical and post-stroke individuals, with 5/5 of the former guessing four or more correctly, and 4/5 of the latter. Notably, S1, who reported only two choices correctly, also had the lowest subjective effort valuation (Table 2), suggesting that this participant was unable to accurately learn from the association phase. A possibility for future research is to employ an additional association phase if the recall phase shows poor performance. However, this would come at the cost of an increased chance of fatigue, which would interfere with subjective effort valuation.

In both subjective effort value measures, we found significant increases when the knee immobilizer was donned. Given the increased energy expenditure with the knee brace, this result was expected [17]. Importantly, this means that the measures of subjective effort value are sensitive enough to detect changes in gait patterns. Further work will be needed to determine whether subjective effort valuation can be used longitudinally throughout recovery.

We did not observe any changes in subjective effort value when comparing people with post-stroke SKG to neurotypical individuals walking with or without the immobilizer. Nominally, subjective effort value was lower in those post-stroke than in neurotypical individuals, contrary to our initial hypothesis. Although the sample size is too small to draw conclusions, it is important to note that neurotypical participants did not have time to acclimate to their new gait pattern, whereas those post-stroke had years. However, because greater cognitive effort is required after stroke for walking [26], we initially predicted that subjective effort value would be higher than in neurotypical individuals in both conditions. Further study is needed to examine whether the effect of stroke on subjective effort value extends beyond changes in biomechanics.

This preliminary study has several limitations. First, our group comparison was conducted in two formats, i.e., treadmill and overground walking. As noted above, greater time for adaptation to knee immobilization would likely have affected the subjective effort value. One post-stroke participant showed a very low subjective effort value (0.34), which would seem to indicate a high readiness to exert effort. It is unclear whether this individual misunderstood the instructions. Instructing the effort choice stage was one of the more challenging parts of the study because it involved discussing probability. Although we collected kinematic data, we did not report it because it was not necessary to determine feasibility. Another limitation of this paradigm is the assumption that all participants will desire to minimize effort. Other factors, such as the desire to challenge oneself, may influence behavior and undermine that assumption. Another factor we did not examine but that likely plays a major role in movement decisions is the fear of falling. Because participants were in a safety harness, the fear of falling was likely greatly reduced compared with a typical situation of being unsupported and alone at home. Future research may wish to delineate these factors.

## VI. Conclusion

This work adapted a novel behavioral economics approach to quantifying subjective effort during walking in people after stroke. We found that the key elements of feasibility were met, albeit with some inter-individual variability. We conclude that this adaptation of subjective effort valuation for people with post-stroke gait impairments is feasible.

## Data Availability

All data produced in the present study are available upon reasonable request to the authors

## Notes

### Competing Interest Statement

The authors have declared no competing interest.

### Funding Statement

This study did not receive any funding

### Author Declarations

The institutional review board at the University of North Carolina at Chapel Hill gave ethical approval for this work. The institutional review board at MetroHealth Medical Center gave ethical approval for this work.

